# Safety of balloon pulmonary angioplasty in chronic thromboembolic pulmonary hypertension: role of the standardized definition of complications

**DOI:** 10.1101/2024.04.19.24306107

**Authors:** F.J. van Leusden, D.P. Staal, M.C.J. van Thor, B.J.M.W. Rensing, J.P. van Kuijk, B.J.M. Mulder, D. van den Heuvel, S. Boerman, K.A. Boomars, J. Peper, J.J. Mager, M.C. Post

## Abstract

Literature reports high complication rates in patients with chronic thromboembolic pulmonary hypertension (CTEPH) who undergo balloon pulmonary angioplasty (BPA), especially in patients with poor pulmonary hemodynamics. Here we describe complications of BPA based on the new definitions.

All patients with CTEPH who completed BPA treatment before September 15th, 2023, were selected from the CTEPH database. Peri-procedural complications were collected and classified according to the 2023 consensus paper on BPA treatment. Complications were analyzed in subgroups of patients with pulmonary vascular resistance (PVR) ≤ or > 6.6 WU and mean pulmonary artery pressure (mPAP) ≤ or > 45 mmHg at first BPA.

In this analysis, 87 patients (63% women; mean age 61.1±14.0 years; 62% on dual PH targeted medical therapy) underwent 426 (mean 4.9±1.6 per patient) BPAs. Only non-severe complications occurred in 14% of BPA treatments; in 47% of the patients; 31% patients had a thoracic complication. The thoracic complications were mild (71%) or moderate (29%). Patients with a PVR >6.6 WU (n=8) underwent more BPA treatments (6.6±1.5 versus 4.6±1.5, p=0.002), had more complications (88% versus 41% of patients, p=0.020), and more thoracic complications (17% vs 7% of BPAs, p=0.013) than patients with PVR ≤6.6 WU. Patients with mPAP >45 mmHg (n=13) also had more BPA-treatments (6.5±1.7 versus 4.6±1.4, p<0.001), more complications (77% versus 44% of patients, p=0.027) and more thoracic complications (14% versus 8% of BPAs, p=0.039) than patients with mPAP ≤45 mmHg.

Complications occurred in 14% of BPAs and were mostly mild. Patients with severe pulmonary hemodynamics suffered more (thoracic) complications.

**What is known?:**

- BPA is an effective treatment in improving pulmonary hemodynamics in patients with CTEPH.
- BPAs are associated with a high complication rate.
- In 2023 a consensus statement was published concerning the definition of complications of BPAs.
- Patients with poor pulmonary hemodynamics may be more susceptible to suffer from complications.

**What does this study add?:**

- For the first time, complications are categorized according to the consensus statement on BPA treatments in patients with CTEPH.
- Thoracic complication rates in patients with severe pulmonary hemodynamics are significantly higher compared to complication rates in patients with lower pulmonary hemodynamic values.

## Introduction

Chronic thromboembolic pulmonary hypertension (CTEPH) is a rare, progressive pathophysiological disorder that is portrayed by persistent pulmonary occlusion, resulting in increased pulmonary vascular resistance (PVR), right heart failure and premature death^1,2^. For patients with proximal obstructive lesions, pulmonary endarterectomy (PEA) is potentially curative and the treatment of choice^1,3,4^. For patients who are ineligible for PEA or for patients with residual pulmonary hypertension (PH) after PEA, balloon pulmonary angioplasty (BPA) offers an alternative, percutaneous interventional approach^1,5–10^. Originally, BPA was associated with severe reperfusion edema, leading to a 30-day mortality rate of 5.5%^5,11^. Mizoguchi et al. introduced BPA as a multiple-staged procedure in 2012, which improved safety considerably and resulted in a prognostic beneficial method for the treatment of CTEPH patients with distal thromboembolic lesions^5,12^.

Peri-procedural complications have been described extensively in the last decade, but until recently no consensus was met concerning the nomenclature of those complications^1,12–14^. In 2023, the first consensus statement on BPA treatment and peri-procedural complications was published by the European Society of Cardiology (ESC)^5^. According to the ESC consensus statement, thoracic complications comprise complications related to the nature of the BPA procedure, the pulmonary arteries, or the underlying pathology, and must be distinguished from non-thoracic complications^5^. Poor pulmonary hemodynamics, based on the PVR and/or mean pulmonary artery pressure (mPAP), have been associated with thoracic complications. Therefore, the ESC guidelines for PH stated that PH-targeted medication should be considered in patients with poor pulmonary hemodynamic status prior to BPA^1,5^.

The aim of this study is to classify complications according to the new nomenclature developed by the ESC and describe the (thoracic) complication rate in patients with severe pulmonary hemodynamics.

## Methods

### Patient selection and data collection

All consecutive patients who were diagnosed with CTEPH and underwent BPA treatment at our hospital from 2015 till the last fully completed BPA procedure in September 2023, were included in the study. For all patients, demographic information (e.g. age, sex, age at diagnosis, medical history), clinical parameters (e.g. World Health Organisation (WHO) functional class, N-terminal prohormone of Brain Natriuretic Peptide (NTproBNP) and 6-minute walking distance (6MWD)), and pulmonary hemodynamics (e.g. mean Right Atrial Pressure (mRAP), mPAP, Pulmonary Artery Wedge Pressure (PAWP), Cardiac Output (CO) and PVR) were collected at the moment of diagnosis, at the first BPA and the last BPA. Information on PH-targeted medication was collected at the time of diagnosis and at the first BPA. All data obtained within six months of the date of diagnosis were considered eligible as baseline characteristics. A right heart catheterization (RHC) was performed during the same session as the first and last BPA. WHO functional class, NTproBNP and 6MWD were considered valid if they were acquired within 3 months of the first BPA or within 3 months after the last BPA. All data were collected and managed using REDCAP electronic data capture tools hosted at the hospital^17,18^. REDCAP is designed to support secure data capture for research studies^17,18^. This study was approved by the local ethical committee of the hospital (MEC-U, Z18.040).

### CTEPH diagnosis and therapeutic management

CTEPH diagnosis and successive assessment of (multimodal) management were established in an expert multidisciplinary team consisting minimally of two PH-physicians (cardiologist and pulmonologist), PEA surgeon, BPA-interventionist, PH-nurse and radiologist, according to the European guidelines for PH^1^. Specific considerations for selecting patients for BPA and/or for PH-targeted medical therapy were described previously in detail^19,20^. Experienced BPA interventionists performed the BPA procedures, following the same principles as published previously by van Thor et al^19^. PH-targeted medical therapy was indicated for CTEPH patients with a WHO functional class of at least II. The time between diagnosis and the first BPA depended on the availability of BPA at the time of diagnosis.

### Peri-procedural complications

Peri-procedural complications were retrospectively collected by evaluating all electronic patient files of patients who underwent BPA. Complications were categorized according to the 2023 consensus statement for BPA^5^. The following were defined as thoracic complications: hemoptysis, vascular injury, lung injury, and ‘other’ thoracic complications. Hemoptysis was defined as mild if less than a hand full of blood was recorded, moderate if more than a hand full of blood was recorded and severe if signs of respiratory failure were present. Patients were considered to have vascular injury if wire perforation, pulmonary artery rupture or pulmonary artery wall dissection was observed during the procedure or additional imaging was performed. Vascular injury was divided into vascular injury with or without hemoptysis, and severity was categorized the same way as it was done for hemoptysis. Pulmonary artery dissection was defined as occlusive or non-occlusive. Lung injury was classified as mild if nasal oxygen was supplied, moderate if non-invasive ventilation was needed, and severe if mechanical ventilation was utilized. Lung injury was classified as immediate if symptoms showed within three hours, -or delayed if more than three hours passed after BPA. Other thoracic complications (e.g. lung infection and pulmonary artery thromboembolism) were also recorded.

Non-thoracic complications constituted of contrast allergy, access site complications, complications related to RHC and contrast nephropathy. Contrast allergy was classified as mild if cutaneous signs of allergy were present, moderate if bronchospasms occurred and severe if signs of anaphylactic shock showed. Complications associated with RHC consisted of (temporary) conduction disturbances, (supra-)ventricular arrythmia, and pericardial tamponade. Contrast nephropathy was classified as acute kidney injury or acute kidney disease, for which dialysis was -or was not needed. Access site complications were recorded if blood transfusion was required or if false aneurysms were present, or if arteriovenous fistulas developed. The details on the definition and therapeutic management of peri-procedural complications can be found in the ESC consensus statement^5^. Mortality, both in-hospital and during follow up were noticed.

### Statistical Analysis

All continuous data are presented as mean ± standard deviation (SD) or as median with interquartile range (IQR), where applicable. All categorical data are presented as number and percentage of total number of patients, or as number and percentage of total number of BPA treatments. Unpaired t-tests were performed to compare complications between subgroups of patients with elevated mPAP or PVR at first BPA. Cut-off values for mPAP >45 mmHg and PVR >6.6 WU were chosen based on previous studies by Wiedenroth et al.^15^ and Jais et al^16^. Statistical tests were two-tailed and were considered significant if p <0.05.

Statistical analysis was performed using R Core Team (2021). R: A language and environment for statistical computing. R Foundation for Statistical Computing, Vienna, Austria. URL https://www.R-project.org/. Version 2023.9.0.463^21^. The following packages were utilized: “tidyverse^22^”, “gtsummary^23^”.

## Results

### Study population and clinical characteristics

In total, 87 patients completed BPA treatments before September 2023 and were included in the present analysis (see figure 1). Their mean age was 61.1±14.0 years and 63% were female. At baseline, 60% of all patients had a WHO functional class of III/IV, mean 6MWD was 380±142 meters and median NTproBNP was 344[108-1788] pg/ml. The baseline mPAP was 38.9±10.0 mmHg and the mean PVR was 6.4±3.6 WU. After diagnosis, 54 (62%) patients received dual PH-targeted therapy. The most prescribed drugs were endothelin receptor antagonists (ERAs) (75%). See table 1 for all baseline characteristics. The median time between diagnosis and the first BPA was 13.8 (IQR 7.4-37.5) months during which the mPAP decreased to 33.1±11.0 mmHg and the PVR to 3.7± 2.3 WU at time of BPA. Other clinical characteristics improved as well (see supplementary table 1). Of the 87 included patients, four patients died before they had completed BPA treatments. One patient died due to acute kidney insufficiency provoked by severe decompensated right heart failure secondary to CTEPH. One patient deceased due to COVID-19 (n=1), one due to Enterococcus faecalis bacteremia resulting in multi-organ failure (n=1) and for one patient the cause of death was unknown.

**Figure 1.**
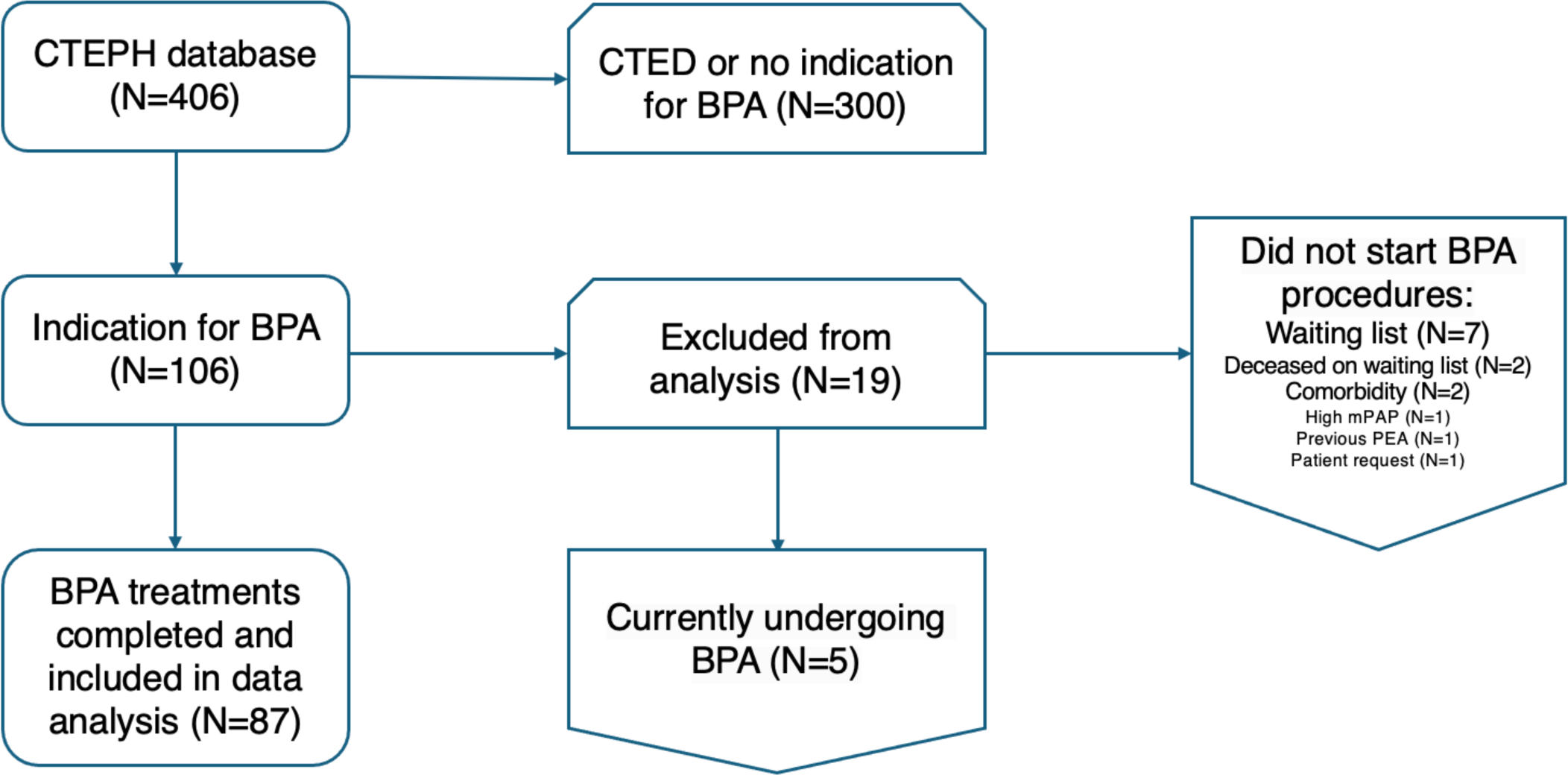
Flowchart of patient inclusion.

**Table 1.** Baseline characteristics of patient population.

|  |  | n (N=87) |  |
| --- | --- | --- | --- |
| **Demographic information** | Sex (female) | 87 | 55 (63.2%) |
|  | Age at diagnosis (years) | 87 | 61.1±14.0 |
|  | BMI >30 kg/m<sup>2</sup> | 69 | 22 (31.9%) |
| **Medical history** | Previous PEA | 87 | 4 (4.6%) |
|  | Systemic hypertension | 85 | 29 (34.1%) |
|  | Diabetes mellitus | 86 | 8 (9.3%) |
|  | CAD | 85 | 3 (3.5%) |
| **Clinical parameters** | WHO FC III/IV | 87 | 52 (59.8%) |
|  | 6MWD (m) | 74 | 380±142 |
|  | NTproBNP (pg/ml)* | 81 | 344[108-1,788] |
| **Pulmonary hemodynamics** | mRAP (mmHg) | 79 | 8.8±5.6 |
|  | mPAP (mmHg) | 86 | 38.9±10.0 |
|  | PAWP (mmHg) | 83 | 10.7±5.0 |
|  | CO (L/Min) | 82 | 5.2±1.9 |
|  | PVR (WU) | 81 | 6.4±3.6 |
| **Anticoagulation type** | DOAC | 87 | 24 (27.6%) |
|  | VKA |  | 63 (72.4%) |
| **PH-targeted medication after the MDT** | No | 87 | 7 (8.0%) |
|  | Mono |  | 25 (28.7%) |
|  | Dual |  | 54 (62.1%) |
|  | Triple |  | 1 (1.1%) |
|  | ERA | 87 | 65 (74.7%) |
|  | Riociguat | 87 | 39 (44.8%) |
|  | PDE5i | 87 | 31 (35.6%) |
|  | Oral prostanoid | 87 | 1 (1.1%) |

BMI = Body Mass Index; BPA = Balloon Pulmonary Angioplasty; CAD = Coronary Artery Disease; CO = Cardiac Output; DOAC = Direct Oral Anticoagulant; ERA = Endothelin Receptor Antagonist; IQR = Interquartile Range; MDT = multidisciplinary team; mPAP = mean Pulmonary Arterial Pressure; mRAP = mean Right Atrial Pressure; n = non-missing; NTproBNP = N-terminal Fragment of Pro-Brain Natriuretic Peptide; PEA = Pulmonary Endarterectomy; PAWP = Pulmonary Arterial Wedge Pressure; PDE5i = Phosphodiesterase-5 Inhibitor; PH = Pulmonary Hypertension; PVR = Pulmonary Vascular Resistance; WHO FC = World Health Organization Functional Class; VKA = Vitamin K Antagonist; 6MWD = 6-minute walking distance. Note: Data are given as mean ± standard deviation or n (%) or \* median [IQR]

### Peri-procedural complications

In total, 426 BPA treatments were performed in 87 patients (4.9±1.6 BPAs per person), of those, 41 (47%) patients suffered at least one complication during the whole treatment period. Most patients had one complication (86%), but some had two (9%), three (2%) or four (2%) complications. During three BPAs more than one complication was recorded, resulting in a total of 59 complications in 57 BPA treatments, and a complication rate of 14% (59/426 BPA treatments). Multiple patients had more than one thoracic complication (38 thoracic complications in 26 patients). Hemoptysis (mostly mild) occurred in 24 (6%), and vascular injury in eight (2%) of the BPA treatments, the latter accompanied by hemoptysis in four cases. Vascular injury constituted of a dissection (n=3), perforation (n=1), rupturing of a sclerotic wall (n=1), or unclear mechanism (n=3). Lung injury occurred after four (1%) BPA treatments and was mostly moderate of nature. Signs of lung injury were immediately present in two cases, while the other two cases were delayed. One patient had to be readmitted after the BPA procedure due to bronchial hyperreactivity, which was classified as any other thoracic complication. Another patient suffered from PH-syncope during the BPA procedure for which cardiopulmonary resuscitation had to be performed for less than 30 seconds.

All signs of contrast allergy were mild. Three complications associated with RHC were supra-ventricular arrhythmia and two complications were temporary conduction disturbances. One patient had signs of temporally acute renal insufficiency, but no dialysis was needed. Six (1%) complications could not be specified according to the new nomenclature for peri-procedural complications: urinary tract infection, bladder retention, collapse due to anxiety, and increased demand for oxygen due to anxiety during the BPA procedure. None of the patients had to be admitted to the intensive care unit and no patients deceased due to complications. All complications are outlined in table 2.

**Table 2.** Overview of peri-procedural complications.

|  | All BPA treatments (N=87) | PVR ≤6.6 (N=64) | PVR >6.6 (N=8) | p-value<sup>1</sup> | mPAP ≤45 (N=69) | mPAP >45 (N=13) | p-value<sup>1</sup> |
| --- | --- | --- | --- | --- | --- | --- | --- |
| **Total BPA count** | 426 | 296 | 53 |  | 317 | 85 |  |
| **BPA count per patient** | 4.9±1.6 | 4.6±1.5 | 6.6±1.5 | 0.002** | 4.6±1.4 | 6.5±1.7 | <0.001*** |
| **Total number of complications** | 59 (13.8%) | 33 (11.1%) | 12 (22.6%) | 0.002** | 40 (12.6%) | 18 (21.2%) | 0.007** |
| **Patients with complications<sup>2</sup>** | 41 (47.1%) | 26 (40.6%) | 7 (87.5%) | 0.020* | 30 (43.5%) | 10 (76.9%) | 0.027* |
| **Patients with thoracic complications<sup>2</sup>** | 27 (31.0%) | 17 (26.6%) | 5 (62.5%) | 0.051 | 19 (27.5%) | 7 (53.8%%) | 0.10 |
| **All thoracic complications** | 38 (8.9%) | 20 (6.8%) | 9 (17.0%) | 0.013* | 25 (7.9%) | 12 (14.1%) | 0.039* |
| Hemoptysis | 24 (5.6%) | 10 (3.4%) | 8 (15.1%) | <0.001*** | 15 (4.7%) | 9 (10.6%) | 0.026* |
| *Mild* | 19 (4.5%) | 10 (3.4%) | 4 (7.5%) | 0.006** | 12 (3.8%) | 7 (8.2%) | 0.093 |
| *Moderate* | 5 (1.2%) | 0 (0.0%) | 4 (7.5%) | <0.001*** | 3 (0.9%) | 2 (2.4%) | 0.064 |
| Vascular injury | 8 (1.9%) | 6 (2.0%) | 0 (0.0%) | 0.38 | 6 (1.9%) | 1 (1.2%) | 0.92 |
| *Mild* | 7 (1.6%) | 5 (1.7%) | 0 (0.0%) | 0.43 | 5 (1.6%) | 1 (1.2%) | 0.97 |
| *Moderate* | 1 (0.2%) | 1 (0.3%) | 0 (0.0%) | 0.76 | 1 (0.3%) | 0 (0.0%) | 0.69 |
| Lung injury | 4 (0.9%) | 2 (0.7%) | 1 (1.9%) | 0.22 | 3 (0.9%) | 1 (1.2%) | 0.62 |
| *Mild* | 1 (0.2%) | 1 (0.3%) | 0 (0.0%) | 0.76 | 1 (0.3%) | 0 (0.0%) | 0.69 |
| *Moderate* | 3 (0.7%) | 1 (0.3%) | 1 (1.9%) | 0.083 | 2 (0.6%) | 1 (1.2%) | 0.41 |
| Other thoracic complications | 2 (0.5%) | 2 (0.7%) | 0 (0.0%) | >0.99 | 1 (0.3%) | 1 (1.2%) | 0.29 |
| **All non-thoracic complications** | 21 (4.9%) | 13 (4.4%) | 3 (5.7%) | 0.48 | 15 (4.7%) | 6 (7.1%) | 0.23 |
| Contrast allergy | 9 (2.1%) | 7 (2.4%) | 0 (0.0%) | 0.43 | 8 (2.5%) | 1 (1.2%) | 0.90 |
| RHC complications | 5 (1.2%) | 3 (1.0%) | 0 (0.0%) | 0.55 | 4 (1.3%) | 1 (1.2%) | 0.81 |
| Contrast nephropathy | 1 (0.2%) | 0 (0.0%) | 0 (0.0%) |  | 0 (0.0%) | 1 (1.2%) |  |
| Access site | 0 (0.0%) | 0 (0.0%) | 0 (0.0%) |  | 0 (0.0%) | 0 (0.0%) |  |
| Other complications | 6 (1.4%) | 3 (1.0%) | 3 (5.7%) | 0.031* | 3 (0.9%) | 3 (3.5%) | 0.12 |
| BPA = Balloon Pulmonary Angioplasty; PVR = Pulmonary Vascular Resistance; RHC = Right Heart CatheterizationNote: Data are given as n (%) of total number of BPA treatments or as mean ± standard deviation |  |  |  |  |  |  |  |
| <sup>1</sup>p <0.05; ** p <0.01; *** p <0.001 |  |  |  |  |  |  |  |

### Peri-procedural complications in patients with mPAP >45 mmHg and PVR >6.6 WU

At the first BPA, eight patients had a PVR >6.6 WU and 13 patients had a mPAP >45 mmHg. Patients with a high PVR underwent 6.6±1.5 BPA treatments, significantly more than patients with a lower PVR (4.6±1.5, p=0.002). Patients with an increased mPAP also underwent significantly more BPA treatments than patients with a mPAP ≤45 mmHg (4.6±1.4 versus 6.5±1.7, p<0.001). Treatments were significantly more often complicated in patients with an increased mPAP or PVR compared to patients with a lower mPAP or PVR (see figure 2). Also, thoracic complications occurred more often in the presence of a high mPAP (14% versus 8%, p=0.039) or high PVR (17% versus 7%, p=0.013) (see figure 3). Finally, ‘other’ types of complications also occurred more often in patients with PVR >6.6 WU (for more details see table 2). Clinical characteristics of patients with an elevated mPAP and PVR are described in supplementary tables 2 and 3. Peri-procedural complications in patients with missing values for mPAP or PVR are described in supplementary tables 4 and 5.

**Figure 2.**
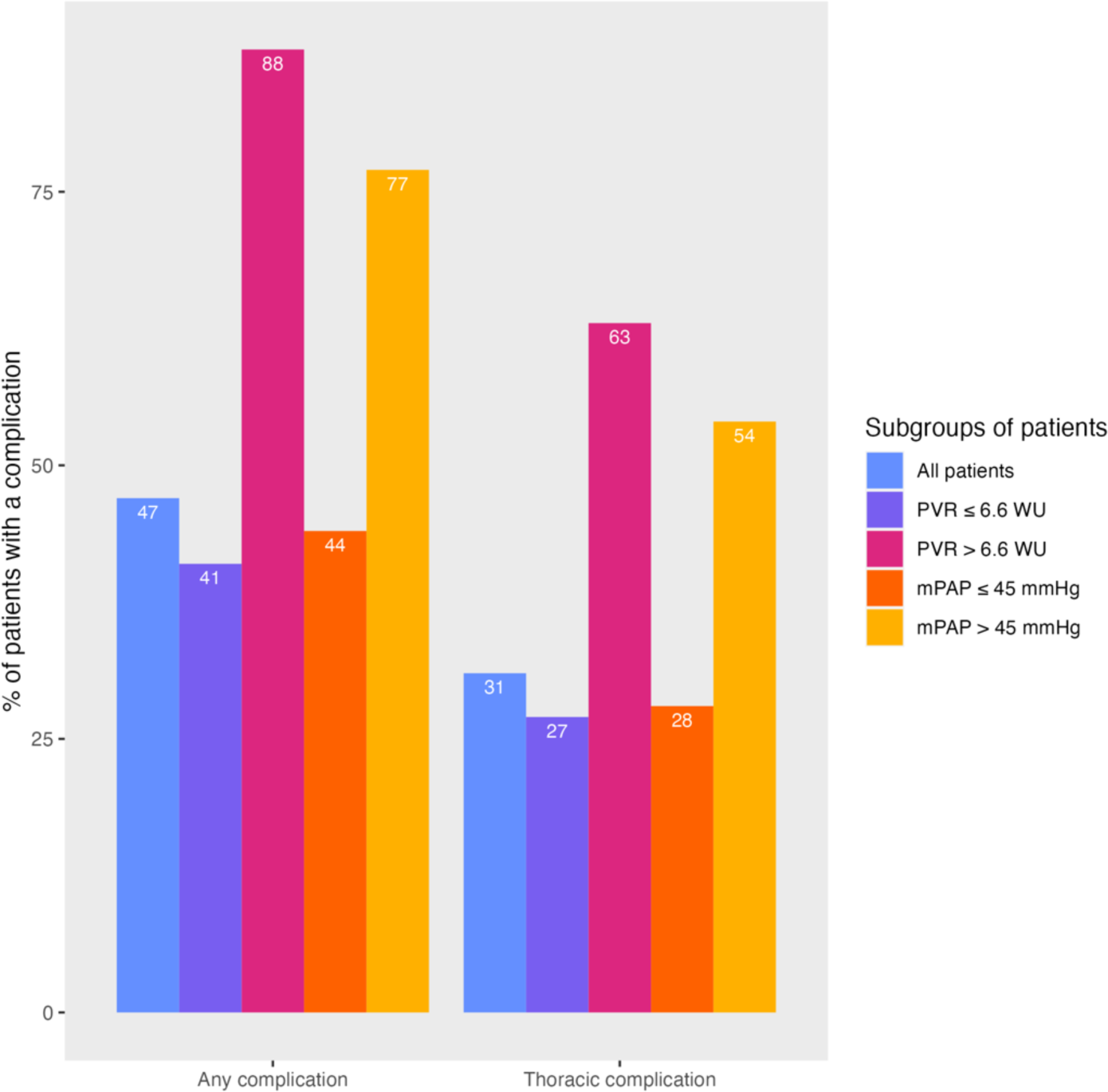
Overview of patients who suffered (thoracic) complications.

**Figure 3.**
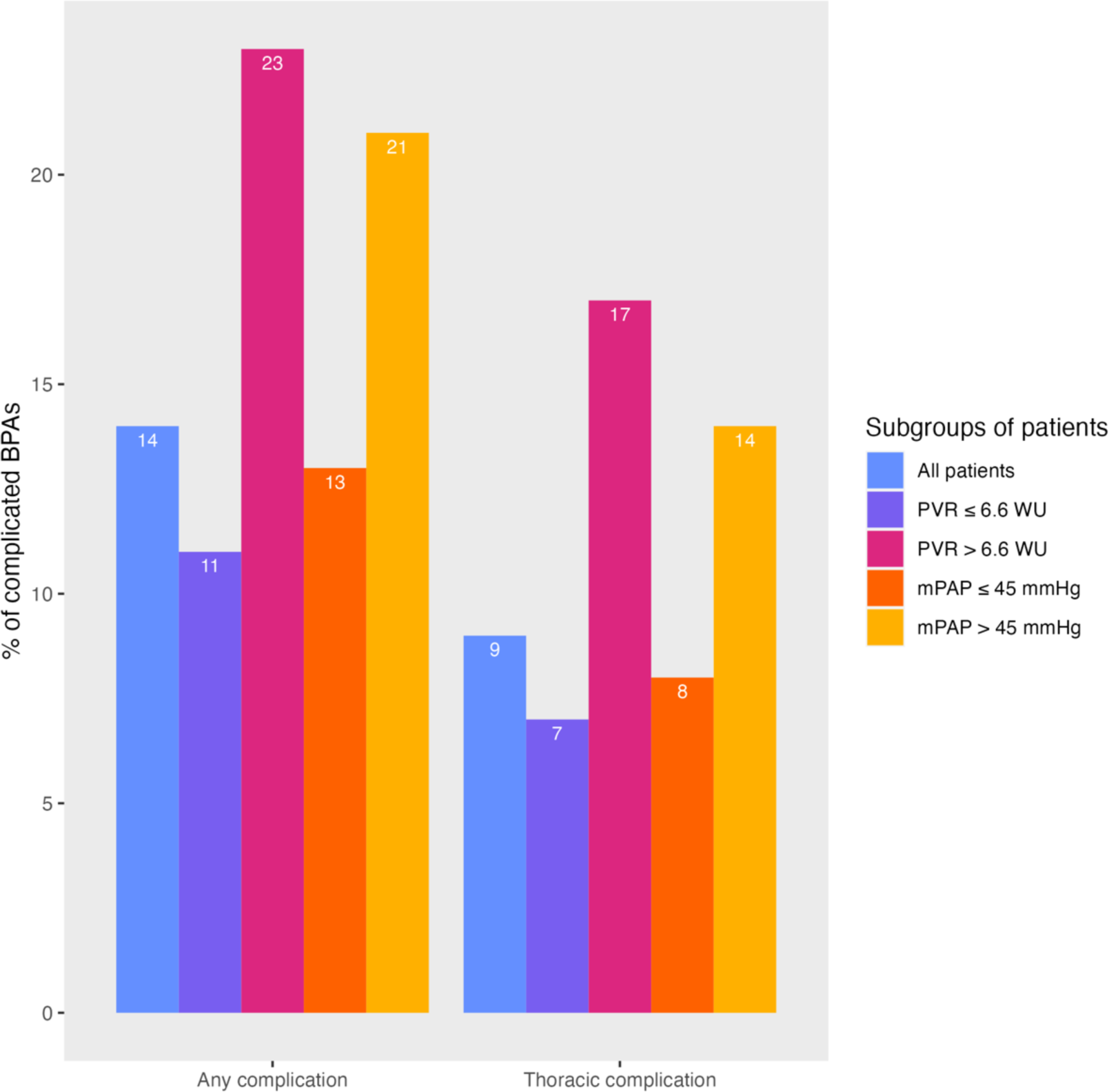
Overview of BPA treatments complicated by (thoracic) complications.

## Discussion

This observational study describes peri-procedural complications according to the recent ESC consensus statement on BPA treatments. The main findings from this analysis are that (1) mild peri-procedural complications are frequently observed and (2) (thoracic) complications, predominantly hemoptysis, are more frequent in patients exhibiting poor pulmonary hemodynamics.

Primarily, the overall prevalence of peri-procedural complications in our study was 14% and 9% for thoracic complications, none of them were severe. The overall complication rate is comparable to the percentages described in other studies, with a range from 11% to 17%^15,24,25,26^. However, in a recent Japanese study by Ito et al.^14^, complications were reported in up to 25% of the BPA treatments. Here, we analyzed mostly inoperable CTEPH patients, in contrast to the Japanese study in which 46% underwent PEA prior to BPA^14^. Previous PEA, and the distinctive thromboembolic phenotype observed in Japanese patients may contribute to the elevated complication rate described by the Japanese investigators compared to the other studies^5,14,25,27,28^. So far, studies on peri-procedural (thoracic) complications were reported prior to the publication of the ESC BPA consensus paper, describing the definition of BPA related complications in detail. Therefore, the comparison of complication rates between different studies should be done with caution^5^. Given the persistent prevalence of (mild) peri-procedural complications globally, the imperative to categorize peri-procedural complications according to the now available nomenclature must be emphasized for future studies^5^.

In a recent study by Wiedenroth et al.^15^ a PVR >6.6 WU was associated with an increased likelihood of thoracic complications, particularly pulmonary vascular perforations. To investigate the cut-off value of 6.6 WU further, we analyzed complications in subgroups of patients with a PVR below and above 6.6 WU. In line with Wiedenroth et al.^15^, we found more (thoracic) complications in the presence of a high PVR compared to a lower PVR (17% versus 7%, p=0.013). The thoracic complication rate, however, was mainly driven by peri-procedural hemoptysis rather than pulmonary vascular injuries. In the ESC consensus statement, hemoptysis is reported as independent thoracic complication as well as symptom of pulmonary artery injury^5^. While hemoptysis is always the consequence of underlying vascular damage, (additional) imaging is needed to identify vascular injury. In this study, not all patients with hemoptysis received imaging after BPA. Therefore, the prevalence of vascular injuries may be underestimated compared to the prevalence of hemoptysis.

Notably, 5 out of the 6 patients (83%) experiencing multiple thoracic complications had a high mPAP and/or high PVR at the first BPA procedure. Among these six patients, four experienced complications of moderate severity. Additionally, 63% of the patients with PVR >6.6 WU suffered thoracic complications versus 27% of the patients with a PVR ≤6.6 WU. In the RACE trial, where patients had a mean baseline PVR of 9.6 WU, 42% of the patients who underwent a BPA had ≥1 treatment-related serious adverse events^16^. These findings suggest that patients with worse pulmonary hemodynamic values are prone to experiencing more, and more serious peri-procedural (thoracic) complications. Accordingly, the imperative to conduct a RHC at the beginning of BPA treatments and the necessity to monitor individuals with worse pulmonary hemodynamic values closely after BPA must be accentuated^5^. Especially, considering that the majority of peri-procedural complications do not cause significant problems when managed adequately^15^.

Finally, the reported absence of severe complications and mortality due to BPA-related complications is unique compared to other publications on peri-procedural complications^14,15,26^. In the ancillary follow-up study after the RACE trial, patients who received riociguat prior to BPA had less (serious) treatment related adverse events than patients who underwent BPA without pre-medical treatment^16^. At our center, PH-targeted therapy was prescribed prior to BPA to more than 90% of the patients. Due to the COVID pandemic, the median duration between diagnosis and the first BPA was long (almost 14 months)^16,24^. Between diagnosis and BPA, a notable improvement was observed in most pulmonary hemodynamic -and clinical parameters, comparable to improvements published in an observational study on riociguat treatment prior to BPA^15^. Interestingly, in 16 patients the PVR decreased to ≤6.6 WU, and in 10 patients the mPAP decreased to ≤45 mmHg. Considering that a high mPAP and/or PVR are associated with (thoracic) complications, the administration of PH-targeted medical therapy may have tempered complication severity and/or influenced peri-procedural complication rate in our study. Future randomized controlled trials are needed to define the role of PH-targeted therapy prior to BPA further. The IMPACT-CTEPH trial is currently ongoing and holds promise in providing insights into the effect of PH-targeted mono therapy versus PH-targeted dual therapy prior to BPA (ClinicalTrials.gov Identifier NCT04780932).

### Limitations

The present analysis must be considered within the context of several limitations. First, the classification of peri-procedural complications relied on clinical experiences rather than on the study of advanced imaging modalities, potentially resulting in some miss classifications. Similarly, complication severity was established based on descriptive measures rather than objective tools. Finally, the single-center character of this analysis must be underscored when generalizing the findings to other settings, particularly considering that PH-targeted therapy was used by most patients prior to BPA.

## Conclusion

This is the first observational study to classify peri-procedural complications according to the ESC consensus statement on BPA treatments. The overall thoracic complication rate is 9%, with a significantly higher rate up to 17% in the presence of severe pulmonary hemodynamics. Despite the retrospective and single-center experience, our analysis establishes a benchmark for future studies on BPA-related complications. The association between increased pulmonary hemodynamics and (thoracic) complications underscores the necessity to investigate the role of PH-targeted therapy preceding BPA further.

## Data Availability

The data that support the findings of this study are available on request from the corresponding author, D.P. Staal. The data are not publicly available due to privacy restrictions.

## Abbreviations

BPA: Balloon Pulmonary Angioplasty
CO: Cardiac Output
CTED: Chronic Thromboembolic Disease
CTEPH: Chronic Thromboembolic Pulmonary Hypertension
ERA: Endothelin Receptor Antagonist
ESC: European Society of Cardiology
IQR: Interquartile Range
MDT: Multi-Disciplinary Team
mPAP: mean Pulmonary Arterial Pressure
mRAP: mean Right Atrial Pressure
NTproBNP: N-terminal prohormone of Brain Natriuretic Peptide
PAWP: Pulmonary Artery Wedge Pressure
PDE5: Phosphodiesterase-5
PEA: Pulmonary Endarterectomy
PH: Pulmonary Hypertension
PVR: Pulmonary Vascular Resistance
RHC: Right Heart Catheterization
SD: Standard Deviation
WHO FC: World Health Organization functional class
WU: Woods Units
6MWD: 6-minute walking distance
95CI: 95% Confidence Interval

## Supplementary Materials

**Supplement table 1:**
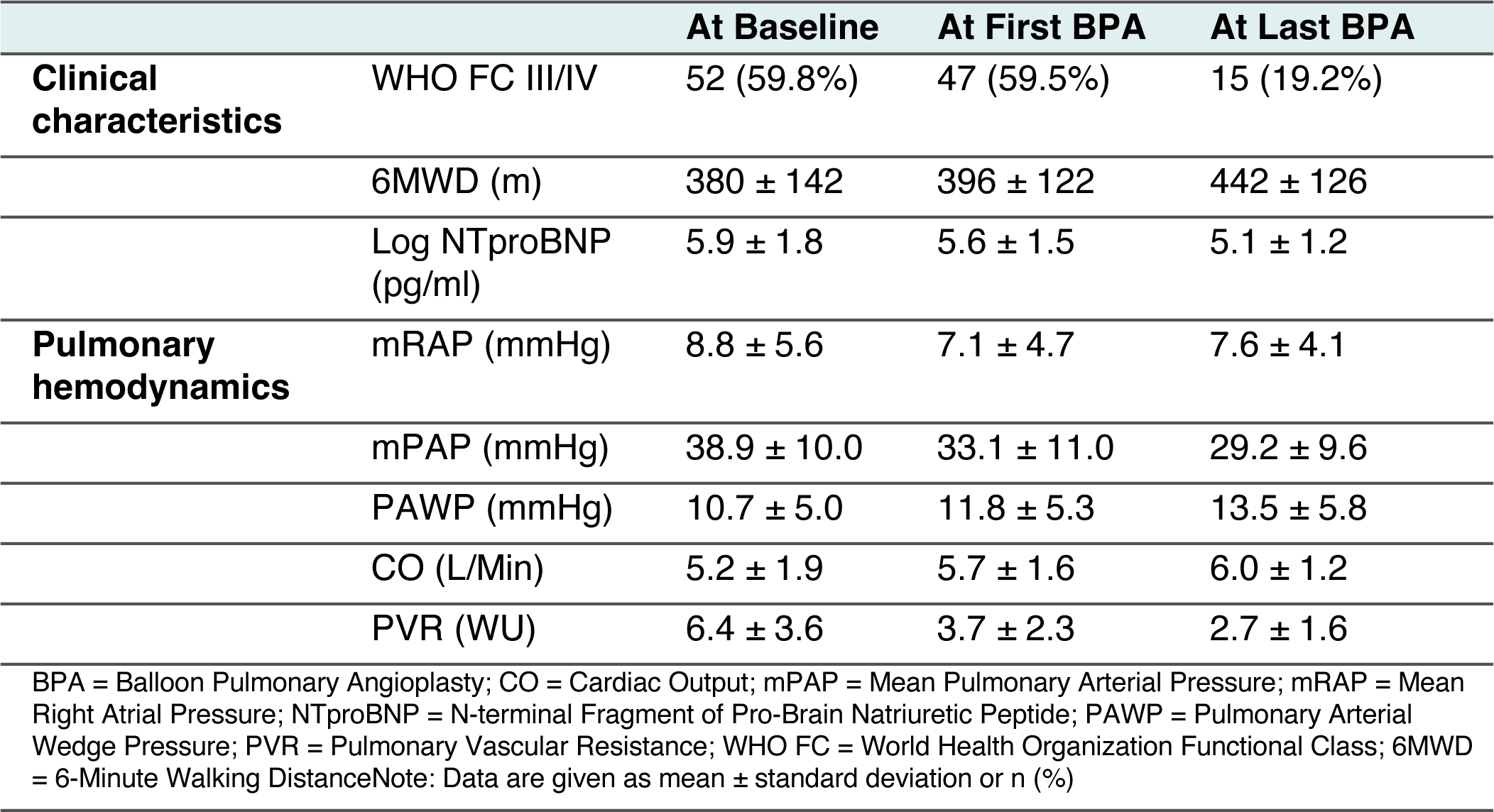
Clinical characteristics at diagnosis, at the first BPA and at the last BPA.

**Supplement table 2:** Clinical characteristics in subgroups of patients with PVR ≤ 6.6 > WU.

|  |  | **PVR $\leq 6.6$**<br>**(N=64)** | **PVR >6.6**<br>**(N=8)** |
| --- | --- | --- | --- |
| **Demographic information** | Sex (female) | 40 (62.5%) | 7 (87.5%) |
| | Age at diagnosis (years) | 61.0 ± 14.3 | 66.5 ± 11.2 |
|  | BMI >30 kg/m<sup>2</sup> | 17 (37.0%) | 0 (0.0%) |
| **Medical history** | Previous PEA | 3 (4.7%) | 0 (0.0%) |
|  | Systemic hypertension | 23 (36.5%) | 3 (37.5%) |
|  | Diabetes mellitus | 5 (7.8%) | 1 (12.5%) |
|  | CAD | 2 (3.2%) | 1 (12.5%) |
| **Clinical parameters** | WHO FC III/IV | 35 (60.3%) | 5 (62.5%) |
| | 6MWD (m) | 403 ± 132 | 380 ± 50 |
|  | NTproBNP (pg/ml), median [IQR] | 168 [99-344] | 2,094 [1,338-2,527] |
| **Pulmonary hemodynamics** | mRAP (mmHg) | 6.6 ± 4.1 | 6.5 ± 3.2 |
| | mPAP (mmHg) | 29.7 ± 8.7 | 46.9 ± 7.6 |
| | PAWP (mmHg) | 12.0 ± 5.4 | 9.1 ± 3.4 |
| | CO (L/Min) | 6.0 ± 1.6 | 4.5 ± 0.9 |
| **BPA** | mean BPA count | 4.6 ± 1.5 | 6.6 ± 1.5 |

BMI = Body Mass Index; BPA = Balloon Pulmonary Angioplasty; CAD = Coronary Artery Disease; CO = Cardiac Output; DOAC = Direct Oral Anticoagulant; IQR = Interquartile Range; mPAP = mean Pulmonary Arterial Pressure; mRAP = mean Right Atrial Pressure; NTproBNP = N-terminal Fragment of Pro-Brain Natriuretic Peptide; PEA = Pulmonary Endarterectomy; PAWP = Pulmonary Arterial Wedge Pressure; PH = Pulmonary Hypertension; PVR = Pulmonary Vascular Resistance; WHO FC = World Health Organization Functional Class; 6MWD = 6-minute walking distance Note: Data are given as mean ± standard deviation or n (%)

**Supplement table 3.**
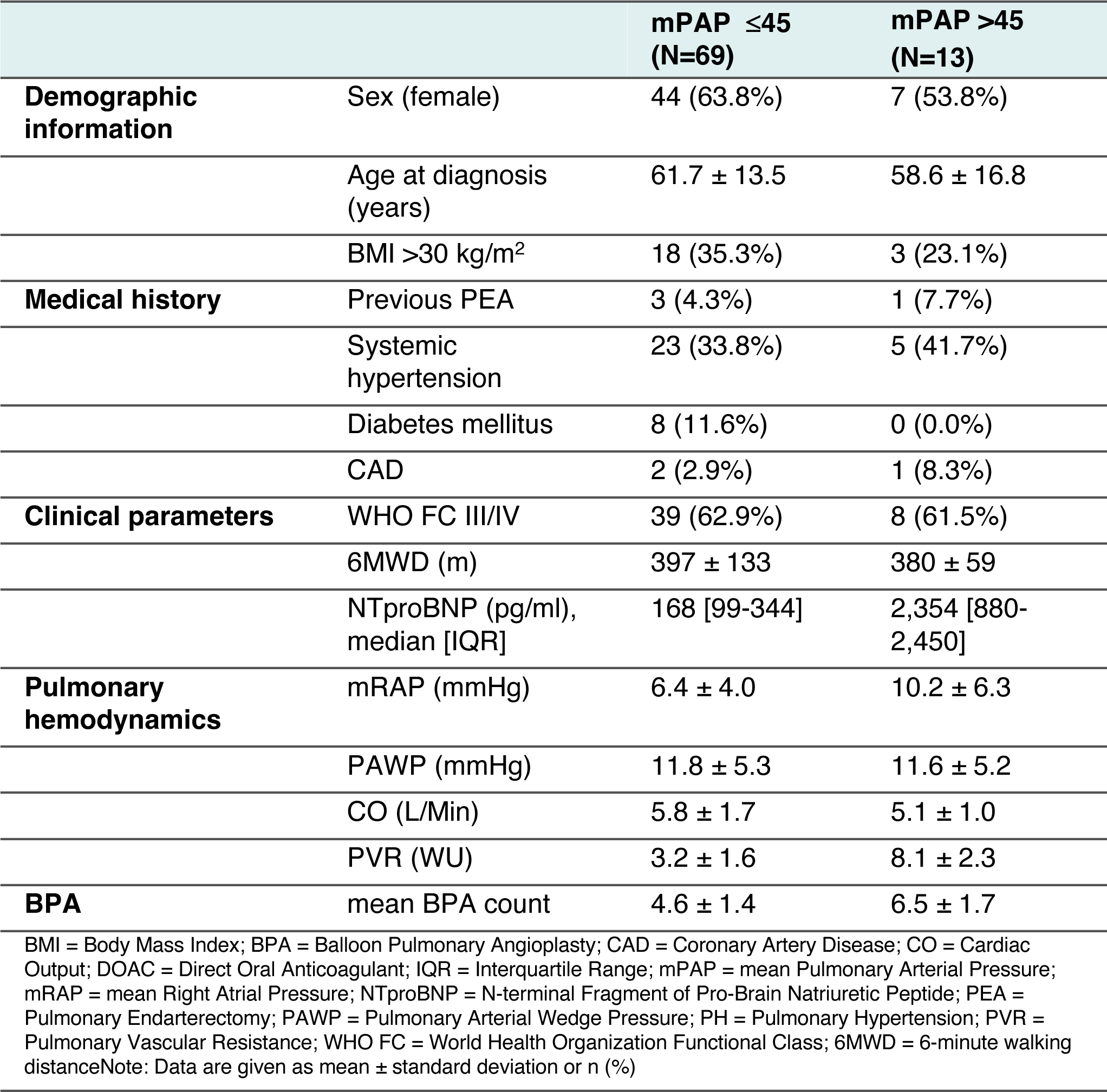
Clinical characteristics of subgroups of patients with mPAP ≤ 45 > mmHg.

**Supplement table 4.** Peri-procedural complications according to PVR status (including missing values for PVR)

|  | **PVR ≤6.6**<br>**(N=64)** | **PVR >6.6**<br>**(N=8)** | **PVR**<br>**missing**<br>**(N=15)** | **p-value<sup>1</sup>** |
| --- | --- | --- | --- | --- |
| **Total BPA count** | 296 | 53 | 77 |  |
| **Mean BPA count** | 4.6 ± 1.5 | 6.6 ± 1.5 | 5.1 ± 1.6 | 0.006** |
| **Total number of complications<sup>2</sup>** | 33 (11.1%) | 12 (22.6%) | 14 (18.2%) | 0.008** |
| **Patients with complications<sup>3</sup>** | 26 (40.6%) | 7 (87.5%) | 8 (53.3%) | 0.031* |
| **Patients with thoracic complications<sup>3</sup>** | 17 (26.6%) | 5 (62.5%) | 5 (33.3%) | 0.14 |
| **Thoracic complications** | 20 (6.8%) | 9 (17.0%) | 9 (11.7%) | 0.048* |
| Hemoptysis during the procedure | 10 (3.4%) | 8 (15.1%) | 6 (7.8%) | 0.001** |
| *Mild* | 10 (3.4%) | 4 (7.5%) | 5 (6.5%) | 0.024* |
| *Moderate* | 0 (0.0%) | 4 (7.5%) | 1 (1.3%) | <0.001*** |
| Vascular injury | 6 (2.0%) | 0 (0.0%) | 2 (2.6%) | 0.57 |
| *Mild* | 5 (1.7%) | 0 (0.0%) | 2 (2.6%) | 0.53 |
| *Moderate* | 1 (0.3%) | 0 (0.0%) | 0 (0.0%) | 0.84 |
| Lung injury | 2 (0.7%) | 1 (1.9%) | 1 (1.3%) | 0.45 |
| *Mild* | 1 (0.3%) | 0 (0.0%) | 0 (0.0%) | 0.84 |
| *Moderate* | 1 (0.3%) | 1 (1.9%) | 1 (1.3%) | 0.21 |
| Other thoracic complications | 2 (0.7%) | 0 (0.0%) | 0 (0.0%) | >0.99 |
| **Non-thoracic complications** | 13 (4.4%) | 3 (5.7%) | 5 (6.5%) | 0.54 |
| Contrast allergy | 7 (2.4%) | 0 (0.0%) | 2 (2.6%) | 0.55 |
| Complications associated with RHC | 3 (1.0%) | 0 (0.0%) | 2 (2.6%) | 0.33 |
| Contrast nephropathy | 0 (0.0%) | 0 (0.0%) | 1 (1.3%) |  |
| Access site | 0 (0.0%) | 0 (0.0%) | 0 (0.0%) |  |
| Others | 3 (1.0%) | 3 (5.7%) | 0 (0.0%) | 0.035* |
| Abbreviations: BPA = Balloon Pulmonary Angioplasty; PVR = Pulmonary Vascular Resistance; RHC = Right Heart CatheterizationNote: Data are given by mean ± standard deviation or as n (%) of number of BPA treatments |  |  |  |  |
| <sup>1</sup>p <0.05; ** p <0.01; *** p <0.001 |  |  |  |  |
| <sup>2</sup>Three BPA treatments were associated with more than one complication |  |  |  |  |
| <sup>3</sup>n (%) of total number of patients |  |  |  |  |

**Supplement table 5.** Peri-procedural complications according to mPAP status (including missing values for mPAP)

|  | mPAP ≤45<br>(N=69) | mPAP >45<br>(N=13) | mPAP<br>missing<br>(N=5) | p-value<sup>1</sup> |
| --- | --- | --- | --- | --- |
| **Total BPA count** | 317 | 85 | 24 |  |
| **Mean BPA count** | 4.6 ± 1.4 | 6.5 ± 1.7 | 4.8 ± 0.8 | 0.003** |
| **Total number of complications<sup>2</sup>** | 40 (12.6%) | 18 (21.2%) | 1 (4.2%) | 0.011* |
| **Patients with complications<sup>3</sup>** | 30 (43.5%) | 10 (76.9%) | 1 (20.0%) | 0.040* |
| **Patients with thoracic complications<sup>3</sup>** | 19 (27.5%) | 7 (53.8%) | 1 (20.0%) | 0.15 |
| **Thoracic complications** | 25 (7.9%) | 12 (14.1%) | 1 (4.2%) | 0.094 |
| Hemoptysis during the procedure | 15 (4.7%) | 9 (10.6%) | 0 (0.0%) | 0.045* |
| *Mild* | 12 (3.8%) | 7 (8.2%) | 0 (0.0%) | 0.14 |
| *Moderate* | 3 (0.9%) | 2 (2.4%) | 0 (0.0%) | 0.14 |
| Vascular injury | 6 (1.9%) | 1 (1.2%) | 4 (4.2%) | 0.69 |
| *Mild* | 5 (1.6%) | 1 (1.2%) | 4 (4.2%) | 0.60 |
| *Moderate* | 1 (0.3%) | 0 (0.0%) | 0 (0.0%) | 0.88 |
| Lung injury | 3 (0.9%) | 1 (1.2%) | 0 (0.0%) | 0.77 |
| *Mild* | 1 (0.3%) | 0 (0.0%) | 0 (0.0%) | 0.88 |
| *Moderate* | 2 (0.7%) | 1 (1.2%) | 0 (0.0%) | 0.63 |
| Other thoracic complications | 1 (0.3%) | 1 (1.2%) | 0 (0.0%) | 0.37 |
| **Non-thoracic complications** | 15 (4.7%) | 6 (7.1%) | 0 (0.0%) | 0.26 |
| Contrast allergy | 8 (2.5%) | 1 (1.2%) | 0 (0.0%) | 0.79 |
| Complications associated with RHC | 4 (1.2%) | 1 (1.2%) | 0 (0.0%) | 0.82 |
| Contrast nephropathy | 0 (0.0%) | 1 (1.2%) | 0 (0.0%) |  |
| Access site | 0 (0.0%) | 0 (0.0%) | 0 (0.0%) |  |
| Others | 3 (0.9%) | 3 (3.5%) | 0 (0.0%) | 0.24 |
| Abbreviations: BPA = Balloon Pulmonary Angioplasty; PVR = Pulmonary Vascular Resistance; RHC = Right Heart Catheterization<br>Note: Data are given by mean ± standard deviation or as n (%) of number of BPA treatments |  |  |  |  |
| <sup>1</sup>p <0.05; ** p <0.01; *** p <0.001 |  |  |  |  |
| <sup>2</sup>Three BPA treatments were associated with more than one complication |  |  |  |  |
| <sup>3</sup>n (%) of total number of patients |  |  |  |  |

## References

1. Humbert M, Kovacs G, Hoeper MM, Badagliacca R, Berger RMF, Brida M, Carlsen J, Coats AJS, Escribano-Subias P, Ferrari P, Ferreira DS, Ghofrani HA, Giannakoulas G, Kiely DG, Mayer E, Meszaros G, Nagavci B, Olsson KM, Pepke-Zaba J, Quint JK, Rådegran G, Simonneau G, Sitbon O, Tonia T, Toshner M, Vachiery J-L, Noordegraaf AV, Delcroix M, Rosenkranz S, Group the ESD. 2022 ESC/ERS Guidelines for the diagnosis and treatment of pulmonary hypertension. European Respiratory Journal. 2022. doi:10.1183/13993003.00879-2022.

2. Moser KM, Bioor CM. Pulmonary Vascular Lesions Occurring in Patients With Chronic Major Vessel Thromboembolic Pulmonary Hypertension. Chest. 1993;103:685–692. 10.1378/chest.103.3.685

3. Jenkins D. Pulmonary endarterectomy: the potentially curative treatment for patients with chronic thromboembolic pulmonary hypertension. European Respiratory Review. 2015;24:263–271. DOI: 10.1183/16000617.00000815

4. Delcroix M, Lang I, Pepke-Zaba J, Jansa P, D’Armini AM, Snijder R, Bresser P, Torbicki A, Mellemkjaer S, Lewczuk J, Simkova I, Barberà JA, Perrot M de, Hoeper MM, Gaine S, Speich R, Gomez-Sanchez MA, Kovacs G, Jaïs X, Ambroz D, Treacy C, Morsolini M, Jenkins D, Lindner J, Dartevelle P, Mayer E, Simonneau G. Long-term outcome of patients with chronic thromboembolic pulmonary hypertension. Circulation. 2016;133:859–871. 10.1161/CIRCULATIONAHA.115.016522

5. Lang IM, Andreassen AK, Andersen A, Bouvaist H, Coghlan G, Escribano-Subias P, Jansa P, Kopec G, Kurzyna M, Matsubara H, Meyer BC, Palazzini M, Post MC, Pruszczyk P, Räber L, Roik M, Rosenkranz S, Wiedenroth CB, Redlin-Werle C, Brenot P. Balloon pulmonary angioplasty for chronic thromboembolic pulmonary hypertension: a clinical consensus statement of the ESC working group on pulmonary circulation and right ventricular function. European Heart Journal. 2023;44:2659–2671. 10.1093/eurheartj/ehad413

6. Delcroix M, Perrot M de, Jaïs X, Jenkins DP, Lang IM, Matsubara H, Meijboom LJ, Quarck R, Simonneau G, Wiedenroth CB, Kim NH. Chronic thromboembolic pulmonary hypertension: Realising the potential of multimodal management. The Lancet Respiratory Medicine. 2023;11:836–850. 10.1016/S2213-2600(23)00292-8

7. Sugimura K, Fukumoto Y, Satoh K, Nochioka K, Miura Y, Aoki T, Tatebe S, Miyamichi-Yamamoto S, Shimokawa H. Percutaneous Transluminal Pulmonary Angioplasty Markedly Improves Pulmonary Hemodynamics and Long-Term Prognosis in Patients With Chronic Thromboembolic Pulmonary Hypertension. Circulation Journal. 2012;76:485–488. 10.1253/circj.cj-11-1217

8. Taniguchi Y, Miyagawa K, Nakayama K, Kinutani H, Shinke T, Okada K, Okita Y, Hirata K, Emoto N. Balloon pulmonary angioplasty: An additional treatment option to improve the prognosis of patients with chronic thromboembolic pulmonary hypertension. EuroIntervention. 2014;10:518–525. 10.4244/EIJV10I4A89

9. Sato H, Ota H, Sugimura K, Aoki T, Tatebe S, Miura M, Yamamoto S, Yaoita N, Suzuki H, Satoh K, Takase K, Shimokawa H. Balloon Pulmonary Angioplasty Improves Biventricular Functions and Pulmonary Flow in Chronic Thromboembolic Pulmonary Hypertension. Circulation Journal. 2016;80:1470–1477. 10.1253/circj.cj-15-1187

10. Lang I, Meyer BC, Ogo T, Matsubara H, Kurzyna M, Ghofrani H-A, Mayer E, Brenot P. Balloon pulmonary angioplasty in chronic thromboembolic pulmonary hypertension. European Respiratory Review. 2017;26. doi:10.1183/16000617.0119-2016.

11. Feinstein JA, Goldhaber SZ, Lock JE, Ferndandes SM, Landzberg MJ. Balloon Pulmonary Angioplasty for Treatment of Chronic Thromboembolic Pulmonary Hypertension. Circulation. 2001;103:10–13. 10.1161/01.cir.103.1.10

12. Mizoguchi H, Ogawa A, Munemasa M, Mikouchi H, Ito H, Matsubara H. Refined Balloon Pulmonary Angioplasty for Inoperable Patients with Chronic Thromboembolic Pulmonary Hypertension. Circulation: Cardiovascular Interventions. 2012;5:748–755. 10.1161/CIRCINTERVENTIONS.112.971077

13. Kennedy MK, Kennedy SA, Tan KT, Perrot M de, Bassett P, McInnis MC, Thenganatt J, Donahoe L, Granton J, Mafeld S. Balloon Pulmonary Angioplasty for Chronic Thromboembolic Pulmonary Hypertension: A Systematic Review and Meta-analysis. CardioVascular and Interventional Radiology. 2022;46:5–18. 10.1007/s00270-022-03323-8

14. Ito R, Yamashita J, Ikeda S, Nakajima Y, Kasahara T, Sasaki Y, Suzuki S, Takahashi L, Komatsu I, Murata N, Shimahara Y, Ogino H, Chikamori T. Predictors of procedural complications in balloon pulmonary angioplasty for chronic thromboembolic pulmonary hypertension. Journal of Cardiology. 2023;82:497–503. 10.1016/j.jjcc.2023.06.011

15. Wiedenroth CB, Deissner H, Adameit MSD, Kriechbaum SD, Ghofrani H-A, Breithecker A, Haas M, Roller F, Rolf A, Hamm CW, Mayer E, Guth S, Liebetrau C. Complications of balloon pulmonary angioplasty for inoperable chronic thromboembolic pulmonary hypertension: Impact on the outcome. The Journal of Heart and Lung Transplantation. 2022;41:1086–1094. 10.1016/j.healun.2022.05.002

16. Jaïs X, Brenot P, Bouvaist H, Jevnikar M, Canuet M, Chabanne C, Chaouat A, Cottin V, De Groote P, Favrolt N, Horeau-Langlard D, Magro P, Savale L, Prévot G, Renard S, Sitbon O, Parent F, Trésorier R, Tromeur C, Piedvache C, Grimaldi L, Fadel E, Montani D, Humbert M, Simonneau G. Balloon pulmonary angioplasty versus riociguat for the treatment of inoperable chronic thromboembolic pulmonary hypertension (RACE): a multicentre, phase 3, open-label, randomised controlled trial and ancillary follow-up study. The Lancet Respiratory Medicine. 2022;10:961–971. 10.1016/S2213-2600(22)00214-4

17. Harris PA, Taylor R, Thielke R, Payne J, Gonzalez N, Conde JG. Research electronic data capture (REDCap)a metadata-driven methodology and workflow process for providing translational research informatics support. Journal of Biomedical Informatics. 2009;42:377–381. 10.1016/j.jbi.2008.08.010

18. Harris PA, Taylor R, Minor BL, Elliott V, Fernandez M, O’Neal L, McLeod L, Delacqua G, Delacqua F, Kirby J, Duda SN. The REDCap consortium: Building an international community of software platform partners. Journal of Biomedical Informatics. 2019;95:103208. 10.1016/j.jbi.2019.103208

19. Thor MCJ van, Lely RJ, Braams NJ, Klooster L ten, Beijk MAM, Heijmen RH, Heuvel DAF van den, Rensing BJWM, Snijder RJ, Vonk Noordegraaf A, Nossent EJ, Meijboom LJ, Symersky P, Mager JJ, Bogaard HJ, Post MC. Safety and efficacy of balloon pulmonary angioplasty in chronic thromboembolic pulmonary hypertension in the Netherlands. Netherlands Heart Journal. 2019;28:81–88. 10.1007/s12471-019-01352-6

20. Thor MCJ van, Snijder RJ, Kelder JC, Mager JJ, Post MC. Does combination therapy work in chronic thromboembolic pulmonary hypertension? International Journal of Cardiology Heart & Vasculature. 2020;29:100544. 10.1016/j.ijcha.2020.100544

21. Posit team. RStudio: Integrated development environment for r. Boston, MA: Posit Software, PBC; 2023. Available at http://www.posit.co/.

22. Wickham H, Averick M, Bryan J, Chang W, McGowan L, François R, Grolemund G, Hayes A, Henry L, Hester J, Kuhn M, Pedersen T, Miller E, Bache S, Müller K, Ooms J, Robinson D, Seidel D, Spinu V, Takahashi K, Vaughan D, Wilke C, Woo K, Yutani H. Welcome to the tidyverse. Journal of Open Source Software. 2019;4:1686.

23. Sjoberg Daniel, D, Whiting K, Curry M, Lavery Jessica, A, Larmarange J. Reproducible Summary Tables with the gtsummary Package. The R Journal. 2021;13:570.

24. Kawakami T, Matsubara H, Shinke T, Abe K, Kohsaka S, Hosokawa K, Taniguchi Y, Shimokawahara H, Yamada Y, Kataoka M, Ogawa A, Murata M, Jinzaki M, Hirata K, Tsutsui H, Sato Y, Fukuda K. Balloon pulmonary angioplasty versus riociguat in inoperable chronic thromboembolic pulmonary hypertension (MR BPA): An open-label, randomised controlled trial. The Lancet Respiratory Medicine. 2022;10:949–960. 10.1016/S2213-2600(22)00171-0

25. Andersen A, Hansen JV, Dragsbaek SJ, Maeng M, Andersen MJ, Andersen G, Mellemjkaer S, Ilkjær LB, Nielsen-Kudsk JE. Balloon pulmonary angioplasty for patients with chronic thromboembolic pulmonary hypertension previously operated by pulmonary endarterectomy. Pulmonary Circulation. 2022;12:e12115. 10.1002/pul2.12115

26. Brenot P, Jaïs X, Taniguchi Y, Garcia Alonso C, Gerardin B, Mussot S, Mercier O, Fabre D, Parent F, Jevnikar M, Montani D, Savale L, Sitbon O, Fadel E, Humbert M, Simonneau G. French experience of balloon pulmonary angioplasty for chronic thromboembolic pulmonary hypertension. European Respiratory Journal. 2019;53. doi:10.1183/13993003.02095-2018.

27. Ito R, Yamashita J, Sasaki Y, Ikeda S, Suzuki S, Murata N, Ogino H, Chikamori T. Efficacy and safety of balloon pulmonary angioplasty for residual pulmonary hypertension after pulmonary endarterectomy. International Journal of Cardiology. 2021;334:105–109. 10.1016/j.ijcard.2021.04.013

28. Chausheva S, Naito A, Ogawa A, Seidl V, Winter M-P, Sharma S, Sadushi-Kolici R, Campean I-A, Taghavi S, Moser B, Klepetko W, Ishida K, Matsubara H, Sakao S, Lang IM. Chronic thromboembolic pulmonary hypertension in Austria and Japan. The Journal of Thoracic and Cardiovascular Surgery. 2019;158:604–614.e2. 10.1016/j.jtcvs.2019.01.019

